# Leveraging large language models for systematic reviewing: A case study using HIV medication adherence research

**DOI:** 10.1101/2024.09.18.24313828

**Authors:** M. Naser Lessani, Zhenlong Li, Shan Qiao, Huan Ning, Abhishek Aggarwal, Guangzhe (Frank) Yuan, Atena Pasha, Michael Stirratt, Lori A. J. Scott-Sheldon

**Author notes:** Corresponding author: Shan Qiao.

## Abstract

**Background:** The rapidly accumulating scientific literature in HIV presents a significant challenge in accurately and efficiently assessing the relevant literature. This study explores the potential capabilities of using large language models (LLMs), such as ChatGPT, for selecting relevant studies for a systematic review.

**Method:** Scientific papers were initially obtained from bibliographic database searches using a Boolean search strategy with pre-defined keywords. From 15,839 unique records, three reviewers manually identified 39 relevant papers based on pre-specified inclusion and exclusion criteria. In the ChatGPT experiment, over 10% of records were randomly chosen as the experimental dataset, including the 39 manually identified manuscripts. These unique records (n=1,680) underwent screening via ChatGPT-4 using the same prespecified criteria. Four strategies were employed including standard prompting, i.e., input-output (IO), chain of thought with zero-shot learning (0-CoT), CoT with few-shot learning (FS-CoT), and Majority Voting (which integrates all three promoting strategies). Performance of the models were assessed using recall, F-score, and precision measures.

**Results:** Recall scores (% of true abstracts successfully identified and retrieved by the model from all input data/records) for different ChatGPT configurations were 0.82 (IO), 0.97 (0-CoT), and both the FS-CoT and the Majority Voting prompts achieved a recall score of 1.0. F-scores were 0.34 (IO), 0.29 (0-CoT), 0.39 (FS-CoT), and 0.46 (majority voting). Precision measures were 0.22(IO), 0.17(0-CoT), 0.24(FS-CoT), and 0.30 (Majority Voting). Computational time varied with 2.32, 4.55, 6.44, and 13.30 hours for IO, 0-CoT, FS-CoT, and majority voting,respectively. Processing costs for the 1,680 unique records were approximately $63, $73, $186, and $325, respectively.

**Conclusion:** LLMs, like ChatGPT, are viable for systematic reviews, efficiently identifying studies meeting pre-specified criteria. Greater efficacy was observed when a more sophisticated prompt design was employed, integrating IO, 0-CoT and FS-CoT prompt techniques (i.e., majority voting). LLMs can expedite the study selection process in systematic reviews compared to manual methods, with minimal cost implications.

## 1 Introduction

The scientific literature is growing at a rapid rate, presenting a significant challenge for comprehensive scientific reviews necessary for guiding clinical decision-making or public policy. It is estimated that over 64 million scientific manuscripts have been published since 1996, with the growth rate of newly published papers increasing over time.

Between 2020 and 2022, the number of published manuscripts increased from 4.68 million to 5.14 million, representing an approximate growth rate of 4.89% (Curcic 2023). The number of published manuscripts in public health has been substantial in recent years. For example, BMC Public Health and BMJ Public Health journals continuously publish a large number of manuscripts related to public health topics.

There is growing interest in investigating how large language models (LLMs) can enhance the efficiency of the screening process in systematic reviews, which is traditionally a laborious and time-consuming task. LLMs, including ChatGPT, are attracting attention from both academic and industrial sectors due to their remarkable abilities across diverse fields (Chang et al. 2024). These models excel in addressing a wide range of general topics, delivering responses that often meet or exceed user expectations. Nonetheless, as the focus shifts towards more specialized or narrow subjects, ChatGPT with standard prompting (IO) may struggle to provide accurate and relevant information. A range of techniques can be paired with standard prompts to enhance the models’ performance. These techniques aim to enhance ChatGPT’s performance on specialized tasks are known as prompt engineering (Wei et al. 2022, Trivedi et al. 2022, Yao et al. 2024, Gramopadhye et al. 2024).

Prompt engineering involves strategically designing inputs for LLMs (i.e., ChatGPT) to elicit specific or enhanced responses, effectively fine-tuning the model’s output to meet precise needs or improve task-specific performance. While the adoption of ChatGPT has grown more widespread across various domains, its most effective utilization relies on how users construct their requests or prompts. Therefore, to fully leverage ChatGPT’s capabilities, understanding the art of prompt engineering is a critical step in the interaction with the model to achieve the task goals (Ekin 2023, Giray 2023, Heston and Khun 2023, Liu et al. 2023). Limited reports address experiences of using LLMs in conducting scientific literature reviews (Sami Abdul Malik et al. 2024, Lessani et al. 2023), and there is a dearth of studies that comprehensively evaluate the feasibility and efficacy of employing LLMs in conducting reviews on HIV-related interventions based on empirical evidence from a high-quality evaluation. In addition, no studies to date have explored prompt engineering strategies for reviews in context of HIV.

Accurately and efficiently screening the HIV-relevant scientific literature, especially literature regarding interventions designed to improve patient adherence to HIV antiretroviral therapy (ART) medications, faces even more challenges. First, participants in ART adherence intervention research have diverse demographic backgrounds, comorbid conditions (e.g., substance use, mental health disorders), and marginalized social identities (e.g., racial/ethnic, sexual, and gender minorities). Second, ART adherence interventions encompass many different approaches and multi-level components (e.g., counseling, peer support, text message reminders, HIV stigma reduction, and so forth). Third, the outcomes in HIV medication adherence intervention research vary from behavioral outcomes (ART uptake or use) to clinical outcomes (e.g., CD4 counts and viral load), and they may incorporate diverse measurement tools such as self-reports, pill counts, electronic health records, and drug level assays. These features of ART adherence intervention research make the screening criteria for systematic reviews complex and likely to require careful prompt enginerering strategies.

The primary objectives of the current study were 1) to assess the feasibility of ChatGPT to expedite the screening process for relevant scientific literature while maintaining accuracy, and 2) to compare capabilities of various prompt engineering techniques on HIV-relaed literature screening.

## 2 Methods

### 2.1. Study selection and inclusion criteria

Following the Preferred Reporting Items for Systematic Reviews and Meta-Analyses (PRISMA) guidelines, (Moher et al. 2009) we searched multiple electronic bibliographic databases (e.g., PubMed, Embase, PsycInfo, CINAHL, Web of Science) for the relevant scientific literature published on or after January 1, 2001. The detailed keyword search terms used for each database are reported in supplement Table 1. Three trained research assistants independently screened the titles and abstracts of all identified records based on pre-defined inclusion and exclusion criteria (see Table 1). The full texts of the identified records were further assessed; discrepancies between reviewers were resolved following discussions with the Principal Investigator.

**Table 1.** Selection criteria of literature review.

| Criteria | Inclusion | Exclusion |
|---|---|---|
| Publication type | Published in journals | Conference materials, theses, dissertations, book reviews, reports |
| Study type | Quantitative studies | Literature reviews, commentaries, protocols, qualitative studies, simulations |
| Design | Randomized controlled trials on HIV treatment adherence and retention | Non-randomized, quasi-experimental, studies without control groups, clinical trials on new treatments |
| Participant age | Adults (≥18 years) | Studies with a mean participant age and age <18 |
| Outcomes reported | Clinical outcomes (e.g., CD4, viral load) and objective medication adherence (e.g. pill count) | Studies focused solely on mental health or other indirect outcomes without reporting clinical data |
| Focus of study | Intervention efficacy or effectiveness | Studies on implementation science without efficacy or effectiveness outcomes |

From 15,839 unique records, the reviewers manually selected 581 abstracts based on inclusion and exclusion criteria during title and abstract screening. Following a full text screening of 581 manuscripts, 39 papers were identified as final inclusions. In the ChatGPT experiment, over 10% of records were randomly chosen as the experimental dataset, including the 39 manually identified true papers (n=1,680).

### 2.2. Prompt Techniques

We employed four types of prompt techniques: Input-output (IO), Chain of Thought with Zero-Shot learning technique (0-CoT), Chain of Thought with Few-Shot learning technique (FS-CoT), and Majority Voting (Yao et al. 2024).

The Input-Output (IO) technique refers to a straightforward approach where the prompt explicitly provides the input followed by an example of the desired output (Brown et al. 2020, Yao et al. 2024). This technique helps guide the model by providing a clear example of the expected output for a given input. It’s akin to showing a model a question (input) and then immediately providing the answer (output), this assists the model in understanding the pattern or format it should follow when generating its response (Figure 1a). This method is particularly useful for tasks where there is a direct relationship between the input and the output, such as translation, summarization, or simple question-answering. By giving the model a clear structure of “if you see this (input), then produce that (output),” it can generate accurate and relevant responses more efficiently, with less computation time.

**Figure 1.**
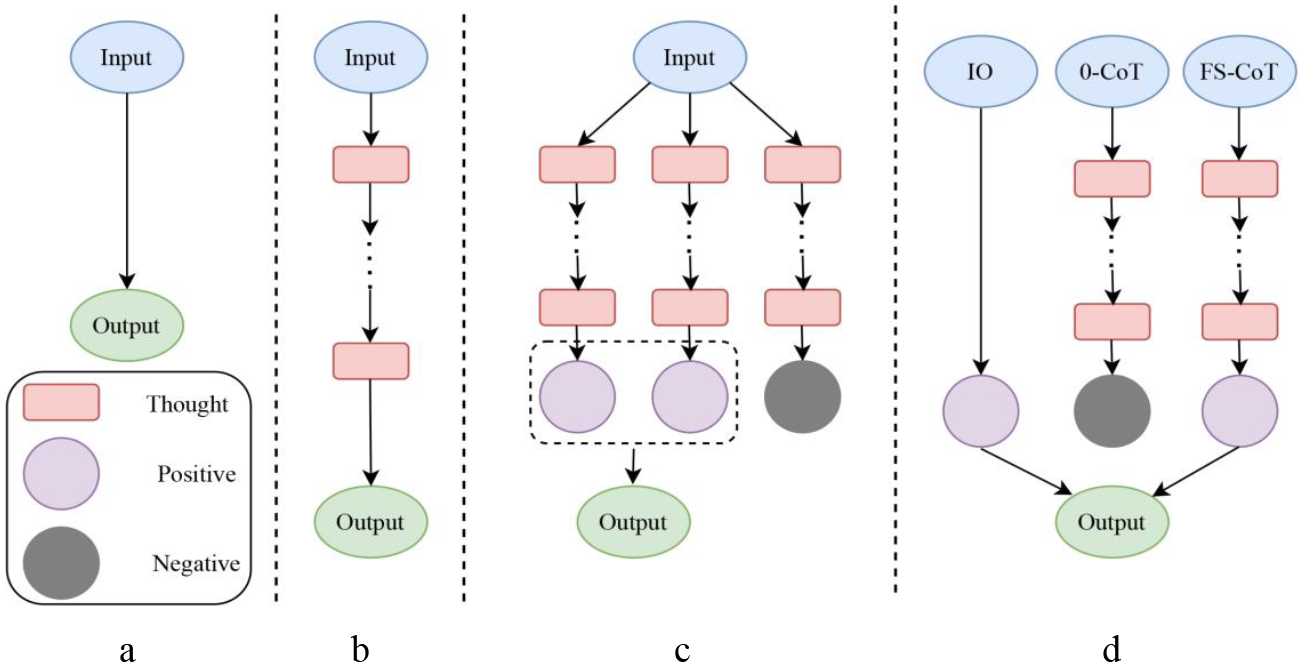
The schematic diagram of three prompt techniques used in this study: a) shows standard prompt (Input and output); b) shows Chain of Thought; c) Majority Voting (Self-Consistency); d) the Majority Voting approach that is used in our experiment. Note, in the figure, labels for thought, positive, and negative indicate the thought process in the model, with outcomes as positive (include the manuscript) or negative (exclude the manuscript). The figure is adopted from (Yao et al. 2024).

0-CoT, on the other hand, is used to solve more complex problems without requiring any specific examples in the prompt (Brown et al. 2020). This approach relies on generating an intermediate, step-by-step reasoning process that leads to the final answer, even when it hasn’t been explicitly trained on similar examples or tasks, as shown in Figure 1b. The “Chain of Thought” part describes how the model internally simulates a logical reasoning process, breaking down the problem into smaller, more manageable parts to conclude. This technique is recommended for challenging questions or problems where direct answers are not immediately obvious, and a reasoned, step-by-step approach can help uncover the solution.

FS-CoT, unlike the zero-shot (0-CoT) approach, attempts to solve problems by utilizing prior examples. These examples serve as a roadmap for the model, illustrating how to break down a problem into smaller, more manageable steps and how to logically progress from those steps to reach a conclusion. In other words, the key idea behind FS-CoT is to prime the model with a few instances where the reasoning process is explicitly laid out, thereby teaching the model the pattern of thought it should emulate when facing a new and similar problem (Wei et al. 2022, Brown et al. 2020). This prompt technique is more advanced than the IO and 0-CoT techniques, mainly leading to a better understanding of the problem and providing a more accurate response to the user’s request.

Majority Voting is an integrative approach used to improve the reliability and accuracy of the model (Wang et al. 2022, Wei et al. 2022). Generally, in this approach, the FS-CoT method is first used to guide the model through a series of reasoning steps for a given problem. After generating multiple responses to the same question, each following an FS-CoT approach but with slight variations in the reasoning process, as illustrated in Figure 1c. Here, each of the generated responses is considered a “vote.” The idea is to analyze these responses and identify the most common answer or conclusion among them. The answer that receives the majority of votes is selected as the final output. This method leverages the principle that, although individual model responses may vary due to nuances in reasoning, the correct answer is more likely to be consistent across multiple attempts.

By aggregating these responses and selecting the most frequent answer, the process aims to filter out outliers and errors, thereby enhancing the overall accuracy and reliability of the model’s output for complex reasoning tasks. The majority voting approach used in this study is illustrated in Figure 1d. Three distinct prompt techniques were applied to each GPT model, and the response receiving the most votes was selected as the final answer.

### 2.3. Model Setting

This study employed the ChatGPT-4 model via API, specifically using the GPT-4-1106-preview version. In the GPT setup, all defined criteria (Table 1) were incorporated into a single input prompt. Figure 2 illustrates the workflow of the model employed, beginning with the title and abstract as input. This is followed by a predefined prompt, to which the input title and abstract are added before submission to GPT. GPT then processes the request and generates a response, which can be a simple ‘include/exclude’ decision. This response also includes reasoning before reaching a decision, employing the Chain of Thought (CoT) concept. It’s important to note that the diagram only shows IO, 0-CoT, and FS-CoT prompt designs. In the Majority Voting approach, normally CoT is iterated several times, but our implementation was specifically based on IO, 0-CoT, and FS-CoT prompts.

**Figure 2.**
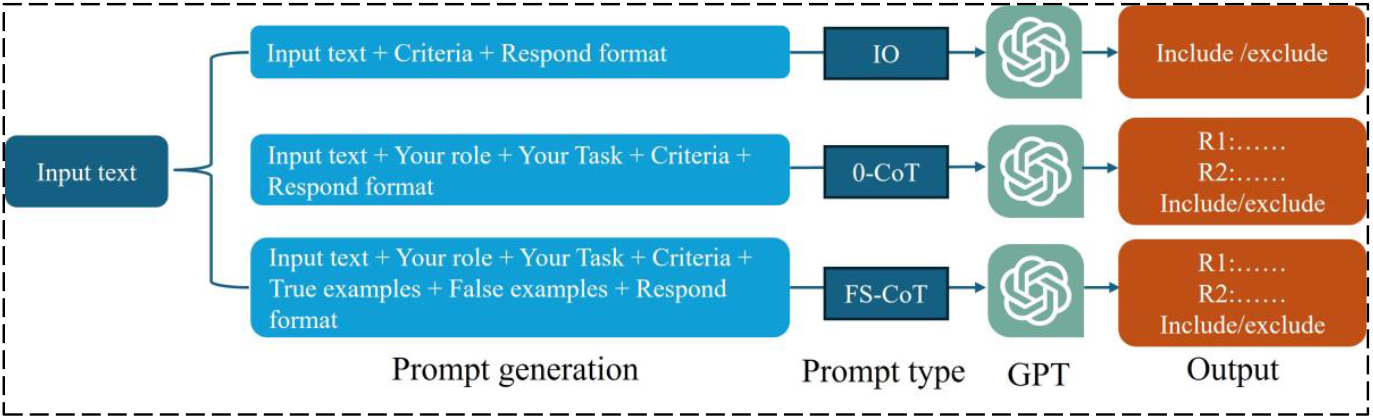
Workflow of the applied model, here ‘R’ stands for reasons.

## 3 Results

### 3.1. Comparison Analysis

The outcomes of the traditional study selection process (in this case, selecting 39 manuscripts out of 1,680 unique records) were regarded as the benchmark for comparison analysis. For comparative analysis, the models’ performance was evaluated using precision, recall, and F-score metrics (Goutte and Gaussier 2005, Derczynski 2016). Precision score measures the proportion of true positives among all predicted positives, essentially, it gauges the accuracy of positive predictions (Equation 1). Recall score, or sensitivity, assesses the proportion of true positives identified from all actual positives, indicating the model’s ability to find all relevant cases (Equation 2). The F-score is the harmonic mean of precision and recall, providing a single metric that balances both precision and recall in one number (Equation 3). In these equations, *“TP”, “FP”*, and *“FN”* stand for true positive, false positive, and false negative, respectively.

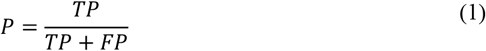

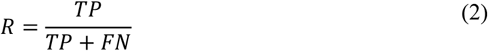

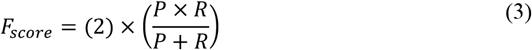

As presented in Table 2, the 0-CoT prompt design yielded the lowest precision score but higher recall and F-scores than IO. FS-CoT and Majority Voting prompt designs recorded the highest recall scores. Majority Voting outperformed all other prompt designs in all assessment parameters, with a notably higher F-score. In this study, we prioritized enhancing the recall score value to ensure that no relevant manuscripts were overlooked during the selection process. Both the FS-CoT and Majority Voting prompts successfully achieved this goal, although FS-CoT selected 30 more abstracts than the Majority Voting strategy. As shown in Table 2, the GPT with IO prompting selected 149 manuscripts out of 1,680, including only 32 relevant (true) manuscripts within this selection. In contrast, using the Majority Voting prompt, GPT selected 131 manuscripts out of 1,680, and all 39 true manuscripts were included in the 131. It is worth noting that only the titles and abstracts of the manuscripts were included in the prompt.

**Table 2.** Comparison of different prompting designs.

| Prompting | Selected manuscripts | Identified relevant manuscripts | Precision | Recall | F-score |
|---|---|---|---|---|---|
| IO | 149 | 32 | 0.215 | 0.820 | 0.340 |
| 0-CoT | 225 | 38 | 0.169 | 0.974 | 0.288 |
| FS-CoT | 161 | 39 | 0.242 | 1.000 | 0.390 |
| Majority Voting | 131 | 39 | 0.297 | 1.000 | 0.459 |

Figure 3 displays the confusion matrices for GPT using four different prompt designs, offering a visual assessment of each model’s classification accuracy by showing the true positives, false positives, true negatives, and false negatives. The GPT model with the IO prompt failed to identify 7 relevant manuscripts, despite having a lower total selection count compared to the 0-CoT and FS-CoT prompts. Meanwhile, the Majority Voting prompt outperformed the others, correctly identifying all relevant manuscripts without any misses and only inaccurately classifying 92 out of 1,680 manuscripts as relevant. This suggests that using the Majority Voting prompt in GPT models can yield more accurate results.

**Figure 3.**
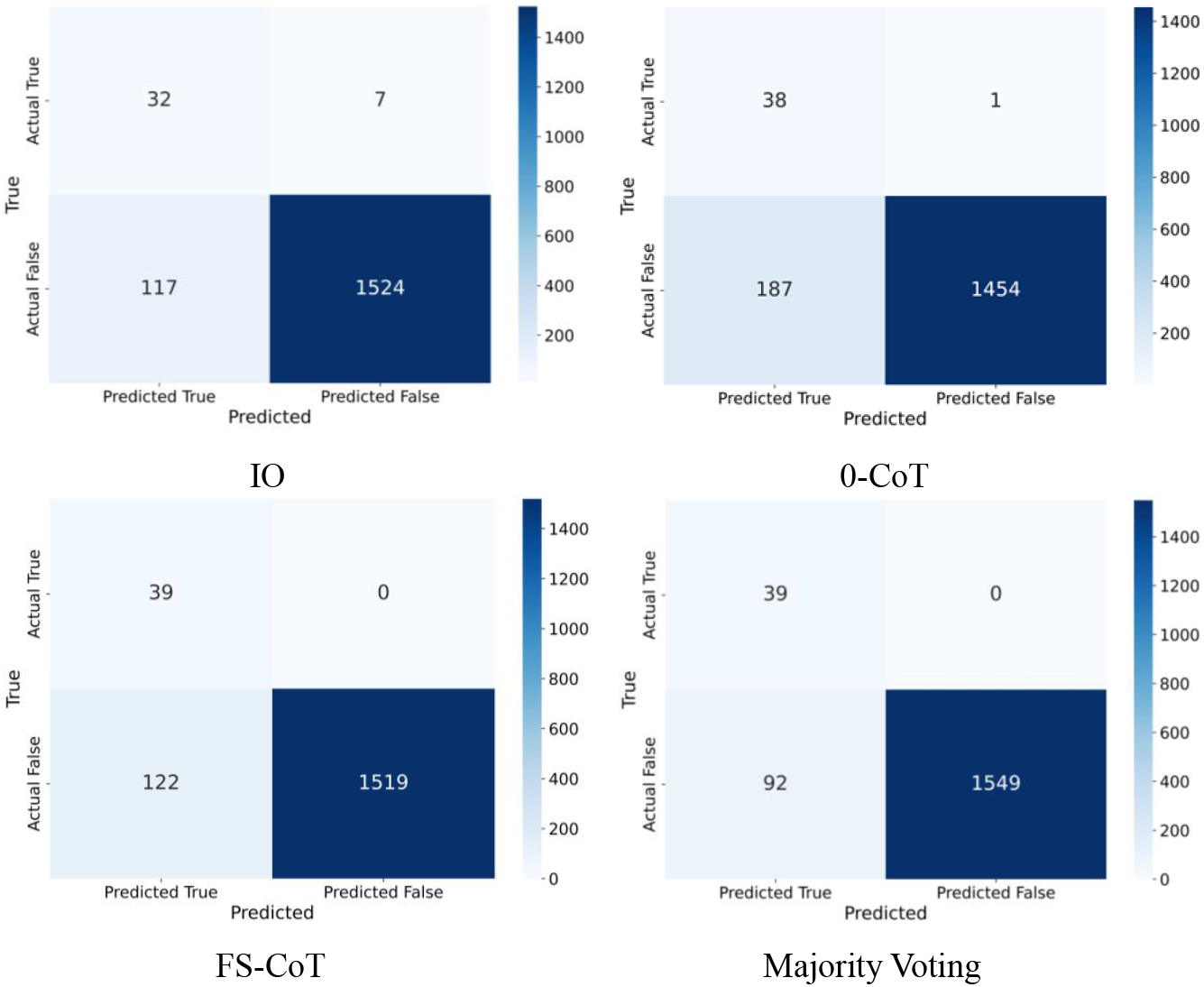
Illustration of the confusion matrix for different prompt designs.

### 3.2. Resource Allocation Analysis

LLMs, including ChatGPT, process text using tokens, which are sequences of characters within the text. As a general guideline, one token typically represents about 4 characters in English text, roughly equivalent to 3/4 of a word (Zhang et al. 2023). The token length in different prompt designs can vary depending on the extent of the text and the specific models used. For instance, The average counts of tokens returned for IO, 0-CoT, and FS-CoT were 1,247, 1,442, and 3,686, respectively. The FS-CoT prompt design used more tokens than the other two prompt designs because it included more detailed explanations, leading to lengthier prompts and more detailed responses, which, in turn, increased the token count. Consequently, the pricing of various GPT models varies based on the number of input and output tokens. As the number of tokens increases, both computation time and associated costs rise. The token count directly depends on the prompt design, for example, the number of tokens in standard prompting is lower, compared to Chain-of-Thought (CoT) prompting.

Table 3 illustrates that the Majority Voting incurs the highest costs among the prompt designs, while IO is the least resource-intensive, as it involves fewer inputs and outputs tokens. The cost and computational time thus are contingent upon the number of tokens processed and the steps required for the model to generate a response. From the perspective of computational time, GPT’s ability to operate independently allows for parallel processing, thereby reducing concerns about computational time. This can greatly accelerate the article selection process, particularly in time-sensitive situations such as public health emergencies and infectious disease outbreaks. The GPT model utilizing the FS-CoT prompt also shows promising results compared to the Majority Voting. The FS-CoT used resources more efficiently, selected fewer manuscripts, but accurately identified all relevant manuscripts without omissions (Figure 4c).

**Table 3.**
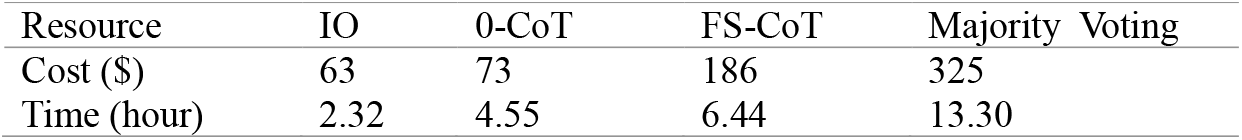
Resource allocation for the model with different prompts.

## 4. Discussion and Limitations

### 4.1. Discussion

Our study demonstrates the feasibility of using ChatGPT for the selection of studies to be used in systematic reviewing. It is also one of the first efforts to compare different prompt techniques in accuracy and efficacy in scientific literature screening. F-scores for various prompt techniques were 0.82 for IO, 0.97 for 0-CoT, and 1.0 for both FS-CoT and Majority Voting. The computational times for these prompting approaches were 2.32, 4.55, 6.44, and 13.30 hours, respectively. The costs to process 1,680 abstracts varied between $63 and $325. It is worth noting that our experiments were completed in March 2024 and based on the GPT-4 model. Both the price and accuracy are likely to change as newer versions of the model are released. For example, according to the GPT website, GPT-4o has reduced costs in half while simultaneously increasing speed (OpenAI 2024). The evidence suggests that ChatGPT could be a promising tool in research synthesis, particularly in reducing the labor-intensive and error-prone task of selecting relevant studies.

Our findings highlight the critical role of prompt strategies in enhancing ChatGPT performance. Prompts serve as the main channel of interaction between users and ChatGPT, guiding the model to produce responses that match the users’ intentions (Ekin 2023, Sahoo et al. 2024). Developing an effective prompt demands domain knowledge, a clear goal and expectation, awareness of the model’s strengths and weaknesses, as well as prompt clarity and specificity (Lo 2023, Ekin 2023). The generation and refinement of the prompts for screening studies (i.e., pre-defined inclusion and exclusion criteria) were based on interdisciplinary communication and discussions. The generated prompts in this study can be applied to other public health topics. Although the specific criteria for including manuscripts may vary across a broad range of public health research topics, the practice of identifying specific inclusion and exclusion criteria is paramount in the research synthesis approach. These include publication period, study design, publication language, target population, and disease conditions. Therefore, our preliminary experiences can inform prompt strategies for screening the scientific literature in other public health domains.

Different prompt strategies produce varying accuracies, measured by F-scores, as well as recall and precision metrics. Based on the F-scores, both FS-CoT and Majority Voting prompts are effective in identifying highly relevant manuscripts from a large dataset. The model utilizing the IO prompt had a significantly lower recall score, suggesting it failed to identify several relevant (true) manuscripts during the selection process. Notably, there is a trade-off between accuracy and costs. For example, although the Majority Voting prompt strategy showed the highest F-score, completing the screening tasks using this strategy took the most hours because this approach requires effort to generate a comprehensive and effective prompt to achieve reliable results. Our study offers evidence to guide decision-making on which prompt strategy to use for article selection in systematic reviews. Thus, scholars can tailor the prompt techniques based on their needs for precision, biases, timelines, and resources.

### 4.2. Limitations

While LLMs (e.g., ChatGPT) have significant capabilities, particularly in text analysis, yet they still come with limitations. Primarily, its performance heavily relies on the design of prompts, posing challenges in the fair evaluation of its effectiveness, particularly in the scientific community where reproducibility and validity are critical. For instance, the model may produce varying responses to identical prompts. Furthermore, the use of LLM may involve associated costs, such as costs posed by using GPT. The estimated cost for our experimental dataset, as shown in Table 3, was not significant. However, for larger datasets and more advanced versions of GPT, the costs increase, potentially becoming a factor to consider. For the Majority Voting in GPT configuration, we considered IO, 0-CoT, and FS-CoT. However, exclusively using the FS-CoT prompt technique could have led to a better performance, albeit at an increased cost for the GPT API.

Several avenues exist for future enhancements. Even though our experiments only considered three prompt techniques in addition to standard prompting (IO), recent advancements have introduced more sophisticated methods, including Graph of Thoughts, Tree of Thoughts, and Everything of Thoughts (Ding et al. 2024). These advanced techniques could significantly improve the performance of the GPT model. More studies are needed to test the accuracy and efficacy of such techniques in LLM in completing other tasks in the literature review, including data extraction, taxonomy development, and findings syntheization. Furthermore, we need to investigate the feasibility and efficacy of using ChatGPT in conducting pilot testing in literature reviews in other disciplines. Additionally, in terms of computation time, we only recorded and estimated the time that was directly spent in running the prompts after refinement of the prompts. We did not include the time for modifying and fine-tuning the prompts. A more detailed cost assessment is needed to estimate and compare the total costs and expenses between the traditional and AI-based approaches.

## 5. Conclusion

Large language models like ChatGPT are viable for systematic reviews, efficiently identifying manuscripts that meet pre-defined inclusion and exclusion criteria. This efficiency is further enhanced when employing a more sophisticated prompting approach, such as the Majority Voting approach designed based on IO, 0-CoT, and FS-CoT prompt techniques in this study. Our findings reveal that these models expedite study selection compared to manual methods, with minimal cost implications. Furthermore, as newer versions of GPT are released, their capabilities and costs improve. For instance, GPT-4o demonstrates real-time applications in audio, vision, and text analysis. This indicates that the newer version of GPT models have the potential not only for study selection but also for extracting knowledge from the selected studies and producing a comprehensive literature review as the final product. Additionally, in the future, the models will be having the capability of identifying research gaps based on the reviewed manuscripts, and will represent a substantial improvement for academics, clinicians, and policymakers.

## Data Availability

All data produced in the present study are available upon reasonable request to the authors

## Appendix 1: Refined prompt that is used for our final experimental analysis

**prompt** = (“Input title and abstract:” + title + abstract + “\n”

“Your role: as a public health professional with nearly 30 years of experience, your expertise spans the entire public health domain. Throughout your career as a public health professor, you have acquired an intricate understanding of the subtle details and differences among various public health topics. Your knowledge is particularly extensive in the area of HIV, where you have significant experience. This background has equipped you with a deep insight into the complexities of public health challenges, especially those related to HIV, enabling you to contribute effectively to the field.”

“Your task: Given your extensive expertise in the public health domain, with a particular focus on HIV, Nature journal has invited you to conduct a literature review on a topic within this area. The specific subject requested for review is HIV medication adherence. You have been tasked with this assignment due to your profound understanding and experience in HIV-related issues.”

“1) Choose the most relevant manuscripts from a collection of abstracts downloaded from online databases, according to specified selection criteria.”

“2) When deciding whether to exclude or include an abstract for further review, you must employ a Chain of Thought and apply your reasoning skills to evaluate it against each selection criterion.”

“3) Your final decision is based on your Chain of Thought and reasoning capabilities.”

“4) Assign a level of relevance, such as strongly relevant, moderately relevant, or weakly relevant.”

“These are the inclusion criteria:

1. The manuscripts must be published in a peer-reviewed journal. Exclude any conference abstracts, conference presentations, conference papers, theses, dissertations, book reviews, or project reports.
2. The studies reported in the manuscripts must be quantitative in nature. Do not include literature reviews [systematic reviews, meta-analyses, scoping reviews, rapid reviews, commentary papers, study protocol papers, modeling simulations, or qualitative studies.]
3. The studies must be intervention studies that utilize a randomized controlled trial [RCT] design. Exclude studies that lack control or comparison groups, employ a quasi-experimental design, only report pre-/post-intervention results, or do not include random assignment.
4. Exclude clinical trials that focus on testing the safety and efficacy of new medicines or new regimens for HIV treatment.
5. The studies must focus on interventions aimed at improving HIV treatment adherence and retention. Do not include studies that only address linkage to care or engagement in HIV treatment without a focus on medication adherence.
6. The studies must involve adult participants [aged 18 years or older at the time of recruitment]. Exclude studies where the mean or median or average age of participants is below 18 years, even if the population is mixed with adults and adolescents. If the study does not mention the age of participants explicitly/clearly, then use your own knowledge and make a decison based on the content of the abstract. For example, check if such studies can be done on participants with age below 18.
7. The studies must report clinical outcomes using continuous variables such as CD4 count, viremia, or viral load, or objective measures of medicine adherence.
8. The studies must report on the efficacy or effectiveness of the interventions.”

“If you’re uncertain about the abstract, then include the article.”

“Your respond should be like this:

Provide your reason and decision against each inclusion and exclusion criteria.

1. First thought, explain why you think this abstract should be included or excluded.”
2. Second thought, provide your reasoning for its inclusion or exclusion.
3. Third thought, explain the rationale behind your decision to include or exclude.”

“Finally your decision, and note that your response should have this format:

{‘Decision’: 1} for inclusion.

{‘Decision’: 0} for exclusion.

{‘Decision’:1} if you’re uncertain about the answer, and I want you to include it.”)

## appendix 2: GPT’s response for one abstract as an example here

Response: 1) First thought: The provided abstract seems to meet some of the inclusion criteria. It is published in a peer-reviewed journal, is based on a quantitative study, and focuses on an intervention aimed at improving HIV treatment adherence.

Second thought: On closer examination, the study detailed in the abstract appears to be an RCT, which is necessary per the inclusion criteria. It compares two different interventions, which indicates the presence of a control or comparison group. The study also involves adult participants and seems to focus on the efficacy of interventions for improving antiretroviral adherence, which is crucial for inclusion.

Third thought: However, the abstract does not explicitly mention the reporting of clinical outcomes using continuous variables such as CD4 count, viremia, or viral load. While the study seems to be related to adherence, which can influence these clinical outcomes, the lack of explicit mention requires assuming this connection based on my knowledge and experience in the field.

Final Decision:

{‘Decision’: 1} - Despite the abstract not explicitly mentioning clinical outcomes, given the direct relationship between antiretroviral adherence and clinical outcomes such as CD4 count and viral load, and based on my expertise in the field, I have decided to include the abstract. The study’s focus on adherence interventions and the use of an RCT design aligns well with the specified selection criteria. If further details are needed, the full article can be reviewed to confirm the reporting of clinical outcomes.

