## Supplement Table 1 for "Leveraging large language models for systematic reviewing: A case study using HIV medication adherence research": Supplement Table 1.docx

*Search string constructed using the search builder in PubMed:* [Advanced Search Results - PubMed (nih.gov)](https://pubmed.ncbi.nlm.nih.gov/advanced/)

**Principles and steps:**

1. Include Main Heading Terms in the [MeSH], treating MeSH entry terms as other regular key terms in the search string
2. Field tag use: we use [tw] for #1 and #2, and use [ti] in #3.
3. We tried different field tag for #1 and #2: text word [tw] first; then Title/abstract [tiab]; then title [ti]
4. Filter use: English Language, human subjects, controlled clinical trial, randomized controlled trial, publication time; review or meta-analysis

#1

"HIV"[Mesh] OR "HIV Infections"[Mesh] OR "Acquired Immunodeficiency Syndrome"[Mesh] OR HIV[tw] OR "HIV Infections"[tw] OR "Acquired Immunodeficiency Syndrome"[tw] OR AIDS[tw] OR "Human Immunodeficiency Viruses"[tw] OR HIV-positive[tw] OR "HIV infect*"[tw] OR "people living with HIV"[tw] OR "people living with HIV/AIDS"[tw] OR "HIV patient*"[tw] OR "highly active antiretroviral therapy"[tw] OR "antiviral therapy"[tw] OR antiretroviral-therapy[tw]

#2

"Treatment Adherence and Compliance"[Mesh] OR "Patient Compliance"[Mesh] OR "Medication Adherence"[Mesh] OR "Retention in Care"[Mesh] OR "Treatment Adherence and Compliance"[tw] OR "Patient Compliance"[tw] OR "Medication Adherence"[tw] OR "Retention in Care"[tw] OR "medication persisten*"[tw] OR "therapeutic adherence"[tw] OR "treatment adherence"[tw] OR "treatment compliance"[tw] OR therapy[tw] OR "therapeutic compliance"[tw] OR "client adherence"[tw] OR "patient adherence"[tw] OR "non-adherent patient"[tw] OR "patient non-adherence"[tw] OR "drug adherence"[tw] OR "medication non-adherence"[tw] OR "care retention"[tw] OR "client compliance"[tw] OR "patient non-compliance"[tw] OR maintain*[tw] OR remain*[tw] OR "viral suppression"[tw] OR viremia[tw] OR "viral load"[tw] OR engagement[tw] OR "care engagement"[tw] OR "re-engagement"[tw]

#3

"Internet-Based Intervention"[Mesh] OR "Psychosocial Intervention"[Mesh] OR "Clinical Trials as Topic"[Mesh] OR "clinical trial"[ti] OR intervention*[ti] OR experiment*[ti] OR promot*[ti] OR maintain*[ti] OR increase*[ti] OR improv*[ti] OR manage*[ti] OR policy[ti] OR law[ti] OR regulation[ti] OR group[ti] OR network[ti] OR clinic*[ti] OR environment[ti] OR communit*[ti] OR "internet-based intervention*"[ti] OR "online intervention*"[ti] OR "internet intervention*"[ti] OR "web-based intervention"[ti] OR "intervention stud*"[ti] OR telephone[ti] OR "text message"[ti] OR smartphone[ti] OR SMS[tiab] OR electronic[ti] OR AI[ti] OR "artificial intelligence"[ti] OR "social media"[ti]

**Searching date: 11/7/2022**

**Database-Specific Search Strings:**

*PubMed*

(("HIV"[MeSH Terms] OR "HIV Infections"[MeSH Terms] OR "Acquired Immunodeficiency Syndrome"[MeSH Terms] OR "HIV"[Text Word] OR "HIV Infections"[Text Word] OR "Acquired Immunodeficiency Syndrome"[Text Word] OR "AIDS"[Text Word] OR "Human Immunodeficiency Viruses"[Text Word] OR "HIV-positive"[Text Word] OR "hiv infect*"[Text Word] OR "people living with HIV"[Text Word] OR "people living with HIV/AIDS"[Text Word] OR "hiv patient*"[Text Word] OR "highly active antiretroviral therapy"[Text Word] OR "antiviral therapy"[Text Word] OR "antiretroviral-therapy"[Text Word]) AND ("Treatment Adherence and Compliance"[MeSH Terms] OR "Patient Compliance"[MeSH Terms] OR "Medication Adherence"[MeSH Terms] OR "Retention in Care"[MeSH Terms] OR "Treatment Adherence and Compliance"[Text Word] OR "Patient Compliance"[Text Word] OR "Medication Adherence"[Text Word] OR "Retention in Care"[Text Word] OR "medication persisten*"[Text Word] OR "therapeutic adherence"[Text Word] OR "treatment adherence"[Text Word] OR "treatment compliance"[Text Word] OR "therapy"[Text Word] OR "therapeutic compliance"[Text Word] OR "client adherence"[Text Word] OR "patient adherence"[Text Word] OR "non-adherent patient"[Text Word] OR "patient non-adherence"[Text Word] OR "drug adherence"[Text Word] OR "medication non-adherence"[Text Word] OR "care retention"[Text Word] OR "client compliance"[Text Word] OR "patient non-compliance"[Text Word] OR "maintain*"[Text Word] OR "remain*"[Text Word] OR "viral suppression"[Text Word] OR "viremia"[Text Word] OR "viral load"[Text Word] OR "engagement"[Text Word] OR "care engagement"[Text Word] OR "re-engagement"[Text Word]) AND ("Internet-Based Intervention"[MeSH Terms] OR "Psychosocial Intervention"[MeSH Terms] OR "Clinical Trials as Topic"[MeSH Terms] OR "clinical trial"[Title] OR "intervention*"[Title] OR "experiment*"[Title] OR "promot*"[Title] OR "maintain*"[Title] OR "increase*"[Title] OR "improv*"[Title] OR "manage*"[Title] OR "policy"[Title] OR "law"[Title] OR "regulation"[Title] OR "group"[Title] OR "network"[Title] OR "clinic*"[Title] OR "environment"[Title] OR "communit*"[Title] OR "internet based intervention*"[Title] OR "online intervention*"[Title] OR "internet intervention*"[Title] OR "web-based intervention"[Title] OR "intervention stud*"[Title] OR "telephone"[Title] OR "text message"[Title] OR "smartphone"[Title] OR "SMS"[Title/Abstract] OR "electronic"[Title] OR "AI"[Title] OR "artificial intelligence"[Title] OR "social media"[Title])) AND ((clinicaltrial[Filter] OR controlledclinicaltrial[Filter] OR evaluationstudy[Filter] OR randomizedcontrolledtrial[Filter]) AND (humans[Filter]) AND (2001/1/1:2022/12/31[pdat]) AND (english[Filter]) AND (alladult[Filter]))

*Embase*

('hiv' OR 'hiv'/exp OR hiv OR 'hiv infections'/exp OR 'hiv infections' OR 'acquired immunodeficiency syndrome'/exp OR 'acquired immunodeficiency syndrome' OR 'aids'/exp OR aids OR 'human immunodeficiency viruses' OR 'hiv positive' OR 'hiv infect*' OR 'people living with hiv'/exp OR 'people living with hiv' OR 'people living with hiv/aids'/exp OR 'people living with hiv/aids' OR 'hiv patient*' OR 'highly active antiretroviral therapy'/exp OR 'highly active antiretroviral therapy' OR 'antiviral therapy'/exp OR 'antiviral therapy' OR 'antiretroviral therapy'/exp OR 'antiretroviral therapy') AND ('treatment adherence and compliance'/exp OR 'treatment adherence and compliance' OR 'patient compliance'/exp OR 'patient compliance' OR 'medication adherence'/exp OR 'medication adherence' OR 'retention in care'/exp OR 'retention in care' OR 'medication persisten*' OR 'therapeutic adherence' OR 'treatment adherence'/exp OR 'treatment adherence' OR 'treatment compliance'/exp OR 'treatment compliance' OR 'therapy'/exp OR therapy OR 'therapeutic compliance' OR 'client adherence' OR 'patient adherence'/exp OR 'patient adherence' OR 'non-adherent patient' OR 'patient non-adherence'/exp OR 'patient non-adherence' OR 'drug adherence'/exp OR 'drug adherence' OR 'medication non-adherence' OR 'care retention'/exp OR 'care retention' OR 'client compliance' OR 'patient non-compliance'/exp OR 'patient non-compliance' OR maintain* OR remain* OR 'viral suppression'/exp OR 'viral suppression' OR 'viremia'/exp OR viremia OR 'viral load'/exp OR 'viral load' OR 'engagement'/exp OR engagement OR 'care engagement' OR 're-engagement') AND ('internet-based intervention'/exp OR 'internet-based intervention' OR 'psychosocial intervention'/exp OR 'psychosocial intervention' OR 'clinical trials as topic'/exp OR 'clinical trials as topic' OR 'clinical trial':ti OR intervention*:ti OR experiment*:ti OR promot*:ti OR maintain*:ti OR increase*:ti OR improv*:ti OR manage*:ti OR policy:ti OR law:ti OR regulation:ti OR group:ti OR network:ti OR clinic*:ti OR environment:ti OR communit*:ti OR 'internet-based intervention*':ti OR 'online intervention*':ti OR 'internet intervention*':ti OR 'web-based intervention':ti OR 'intervention stud*':ti OR telephone:ti OR 'text message':ti OR smartphone:ti OR sms:ti OR electronic:ti OR ai:ti OR 'artificial intelligence':ti OR 'social media':ti) AND ([controlled clinical trial]/lim OR [randomized controlled trial]/lim) AND [2001-2022]/py AND [01-01-2001]/sd NOT [01-01-2023]/sd AND ('clinical trial'/de OR 'controlled clinical trial'/de OR 'controlled study'/de OR 'intervention study'/de OR 'randomized controlled trial'/de) AND [adult]/lim

PsycINFO

("HIV"[Mesh] OR "HIV Infections"[Mesh] OR "Acquired Immunodeficiency Syndrome"[Mesh] OR HIV OR "HIV Infections" OR "Acquired Immunodeficiency Syndrome" OR AIDS OR "Human Immunodeficiency Viruses" OR HIV-positive OR "HIV infect*" OR "people living with HIV" OR "people living with HIV/AIDS" OR "HIV patient*" OR "highly active antiretroviral therapy" OR "antiviral therapy" OR antiretroviral-therapy)

("Treatment Adherence and Compliance"[Mesh] OR "Patient Compliance"[Mesh] OR "Medication Adherence"[Mesh] OR "Retention in Care"[Mesh] OR "Treatment Adherence and Compliance" OR "Patient Compliance" OR "Medication Adherence" OR "Retention in Care" OR "medication persisten*" OR "therapeutic adherence" OR "treatment adherence" OR "treatment compliance" OR therapy OR "therapeutic compliance" OR "client adherence" OR "patient adherence" OR "non-adherent patient" OR "patient non-adherence" OR "drug adherence" OR "medication non-adherence" OR "care retention" OR "client compliance" OR "patient non-compliance" OR maintain* OR remain* OR "viral suppression" OR viremia OR "viral load" OR engagement OR "care engagement" OR "re-engagement")

("Internet-Based Intervention"[Mesh] OR "Psychosocial Intervention"[Mesh] OR "Clinical Trials as Topic"[Mesh] OR "clinical trial" OR intervention* OR experiment* OR promot* OR maintain* OR increase* OR improv* OR manage* OR policy OR law OR regulation OR group OR network OR clinic* OR environment OR communit* OR "internet-based intervention*" OR "online intervention*" OR "internet intervention*" OR "web-based intervention" OR "intervention stud*" OR telephone OR "text message" OR smartphone OR SMS OR electronic OR "artificial intelligence" OR "social media")

**Specifications**

- Removed AI from last string
- All text for first two
- Title for the last

#### Filters[X](https://web.s.ebscohost.com/ehost/breadbox/remove?item=limiter_PY&sid=e60811ab-339e-4782-a466-b8a213b9a3cc%40redis&vid=2)

- Publication Year: 2001-2022
- English
- Age Groups: Adulthood (18 yrs & older)
- Population Group: Human

*CINAHL*

("HIV"[Mesh] OR "HIV Infections"[Mesh] OR "Acquired Immunodeficiency Syndrome"[Mesh] OR HIV OR "HIV Infections" OR "Acquired Immunodeficiency Syndrome" OR AIDS OR "Human Immunodeficiency Viruses" OR HIV-positive OR "HIV infect*" OR "people living with HIV" OR "people living with HIV/AIDS" OR "HIV patient*" OR "highly active antiretroviral therapy" OR "antiviral therapy" OR antiretroviral-therapy)

("Treatment Adherence and Compliance"[Mesh] OR "Patient Compliance"[Mesh] OR "Medication Adherence"[Mesh] OR "Retention in Care"[Mesh] OR "Treatment Adherence and Compliance" OR "Patient Compliance" OR "Medication Adherence" OR "Retention in Care" OR "medication persisten*" OR "therapeutic adherence" OR "treatment adherence" OR "treatment compliance" OR therapy OR "therapeutic compliance" OR "client adherence" OR "patient adherence" OR "non-adherent patient" OR "patient non-adherence" OR "drug adherence" OR "medication non-adherence" OR "care retention" OR "client compliance" OR "patient non-compliance" OR maintain* OR remain* OR "viral suppression" OR viremia OR "viral load" OR engagement OR "care engagement" OR "re-engagement")

("Internet-Based Intervention"[Mesh] OR "Psychosocial Intervention"[Mesh] OR "Clinical Trials as Topic"[Mesh] OR "clinical trial" OR intervention* OR experiment* OR promot* OR maintain* OR increase* OR improv* OR manage* OR policy OR law OR regulation OR group OR network OR clinic* OR environment OR communit* OR "internet-based intervention*" OR "online intervention*" OR "internet intervention*" OR "web-based intervention" OR "intervention stud*" OR telephone OR "text message" OR smartphone OR SMS OR electronic OR "artificial intelligence" OR "social media")

**Specifications**

- Removed AI from last string
- All text for first two
- Title for the last

#### Filters

- Full Text
- Published Date: 20010101-20221231
- English Language
- Human
- Language: English
- Age Groups: All Adult

*Eric*

("HIV"[Mesh] OR "HIV Infections"[Mesh] OR "Acquired Immunodeficiency Syndrome"[Mesh] OR HIV OR "HIV Infections" OR "Acquired Immunodeficiency Syndrome" OR AIDS OR "Human Immunodeficiency Viruses" OR HIV-positive OR "HIV infect*" OR "people living with HIV" OR "people living with HIV/AIDS" OR "HIV patient*" OR "highly active antiretroviral therapy" OR "antiviral therapy" OR antiretroviral-therapy)

("Treatment Adherence and Compliance"[Mesh] OR "Patient Compliance"[Mesh] OR "Medication Adherence"[Mesh] OR "Retention in Care"[Mesh] OR "Treatment Adherence and Compliance" OR "Patient Compliance" OR "Medication Adherence" OR "Retention in Care" OR "medication persisten*" OR "therapeutic adherence" OR "treatment adherence" OR "treatment compliance" OR therapy OR "therapeutic compliance" OR "client adherence" OR "patient adherence" OR "non-adherent patient" OR "patient non-adherence" OR "drug adherence" OR "medication non-adherence" OR "care retention" OR "client compliance" OR "patient non-compliance" OR maintain* OR remain* OR "viral suppression" OR viremia OR "viral load" OR engagement OR "care engagement" OR "re-engagement")

("Internet-Based Intervention"[Mesh] OR "Psychosocial Intervention"[Mesh] OR "Clinical Trials as Topic"[Mesh] OR "clinical trial" OR intervention* OR experiment* OR promot* OR maintain* OR increase* OR improv* OR manage* OR policy OR law OR regulation OR group OR network OR clinic* OR environment OR communit* OR "internet-based intervention*" OR "online intervention*" OR "internet intervention*" OR "web-based intervention" OR "intervention stud*" OR telephone OR "text message" OR smartphone OR SMS OR electronic OR "artificial intelligence" OR "social media")

**Specifications**

- Removed AI from last string
- All text for first two
- Title for the last

#### Filters

- Date Published: 20010101-20221231
- Language: English

*ProQuest Dissertation Thesis and Global*

noft((HIV OR "Acquired Immunodeficiency Syndrome" OR AIDS OR "Human Immunodeficiency Viruses" OR HIV-positive OR "HIV infect*" OR "people living with HIV" OR "people living with HIV/AIDS" OR "HIV patient*" OR "highly active antiretroviral therapy" OR "antiviral therapy" OR antiretroviral-therapy)) AND noft(("Treatment Adherence and Compliance" OR "Patient Compliance" OR "Medication Adherence" OR "Retention in Care" OR "Treatment Adherence and Compliance" OR "Patient Compliance" OR "medication persisten*" OR "therapeutic adherence" OR "treatment adherence" OR "treatment compliance" OR therapy OR "therapeutic compliance" OR "client adherence" OR "patient adherence" OR "non-adherent patient" OR "patient non-adherence" OR "drug adherence" OR "medication non-adherence" OR "care retention" OR "client compliance" OR "patient non-compliance" OR maintain* OR remain* OR "viral suppression" OR viremia OR "viral load" OR engagement OR "care engagement" OR "re-engagement")) AND title(("Internet-based Intervention" OR "Psychosocial Intervention" OR "Clinical Trials as Topic" OR "clinical trial" OR intervention* OR experiment* OR promot* OR maintain* OR increase* OR improv* OR manage* OR policy OR law OR regulation OR group OR network OR clinic* OR environment OR communit* OR "online intervention*" OR "internet intervention*" OR "web-based intervention" OR "intervention stud*" OR telephone OR "text message" OR smartphone OR SMS OR electronic OR "artificial intelligence" OR AI OR "social media"))

**Filters:** Years and language filter applied

*Web of Science*

((TI=(("HIV"[Mesh] OR "HIV Infections"[Mesh] OR "Acquired Immunodeficiency Syndrome"[Mesh] OR HIV OR "HIV Infections" OR "Acquired Immunodeficiency Syndrome" OR AIDS OR "Human Immunodeficiency Viruses" OR HIV-positive OR "HIV infect*" OR "people living with HIV" OR "people living with HIV/AIDS" OR "HIV patient*" OR "highly active antiretroviral therapy" OR "antiviral therapy" OR antiretroviral-therapy))) AND TI=(("Treatment Adherence and Compliance"[Mesh] OR "Patient Compliance"[Mesh] OR "Medication Adherence"[Mesh] OR "Retention in Care"[Mesh] OR "Treatment Adherence and Compliance" OR "Patient Compliance" OR "Medication Adherence" OR "Retention in Care" OR "medication persisten*" OR "therapeutic adherence" OR "treatment adherence" OR "treatment compliance" OR therapy OR "therapeutic compliance" OR "client adherence" OR "patient adherence" OR "non-adherent patient" OR "patient non-adherence" OR "drug adherence" OR "medication non-adherence" OR "care retention" OR "client compliance" OR "patient non-compliance" OR maintain* OR remain* OR "viral suppression" OR viremia OR "viral load" OR engagement OR "care engagement" OR "re-engagement"))) AND TI=(("Internet-Based Intervention"[Mesh] OR "Psychosocial Intervention"[Mesh] OR "Clinical Trials as Topic"[Mesh] OR "clinical trial" OR intervention* OR experiment* OR promot* OR maintain* OR increase* OR improv* OR manage* OR policy OR law OR regulation OR group OR network OR clinic* OR environment OR communit* OR "internet-based intervention*" OR "online intervention*" OR "internet intervention*" OR "web-based intervention" OR "intervention stud*" OR telephone OR "text message" OR smartphone OR SMS OR electronic OR AI OR "artificial intelligence" OR "social media"))

*Cochrane Library*

**Title abstract – title abstract – record title**

(HIV OR "Acquired Immunodeficiency Syndrome" OR AIDS OR "Human Immunodeficiency Viruses" OR HIV-positive OR "HIV infect*" OR "people living with HIV" OR "people living with HIV/AIDS" OR "HIV patient*" OR "highly active antiretroviral therapy" OR "antiviral therapy" OR antiretroviral-therapy)

("Treatment Adherence and Compliance" OR "Patient Compliance" OR "Medication Adherence" OR "Retention in Care" OR "Treatment Adherence and Compliance" OR "Patient Compliance" OR "medication persisten*" OR "therapeutic adherence" OR "treatment adherence" OR "treatment compliance" OR therapy OR "therapeutic compliance" OR "client adherence" OR "patient adherence" OR "non-adherent patient" OR "patient non-adherence" OR "drug adherence" OR "medication non-adherence" OR "care retention" OR "client compliance" OR "patient non-compliance" OR maintain* OR remain* OR "viral suppression" OR viremia OR "viral load" OR engagement OR "care engagement" OR "re-engagement")

("Internet-based Intervention" OR "Psychosocial Intervention" OR "Clinical Trials as Topic" OR "clinical trial" OR intervention* OR experiment* OR promot* OR maintain* OR increase* OR improv* OR manage* OR policy OR law OR regulation OR group OR network OR clinic* OR environment OR communit* OR "online intervention*" OR "internet intervention*" OR "web-based intervention" OR "intervention stud*" OR telephone OR "text message" OR smartphone OR SMS OR electronic OR "artificial intelligence" OR AI OR "social media")

**Filter:** Added date filter
